# A “vulnerable” space: Health professions education scholars’ reflections on AI-use disclosure

**DOI:** 10.64898/2026.10.01.26364502

**Authors:** L. Lingard, L. A. Maggio, M. Ans, E. Driessen

## Abstract

**Background:** AI use is growing by HPE scholars, and disclosure of that use is required by scientific ethics and journal policy. In spite of growing unease about the meaning, and meaningfulness, of AI-use disclosures in published manuscripts, little is known about how authors represent their AI use in published disclosure statements. Without such understanding, we are ill-equipped to redirect disclosure practices for increased sufficiency and sincerity, undermining efforts towards transparency in the era of AI-supported scholarship

**Methods:** In this descriptive qualitative study, 18 HPE researchers with experience disclosing generative AI use in publications from 2025-2026 were interviewed using Zoom. Data collection and thematic analysis proceeded iteratively, exploring participants’ experiences with AI use, their disclosure decisions, and evolving disclosure practices.

**Results:** Participants reported a wide range of experience writing AI-use disclosures (from 1-16 disclosures/participant); in total 31 published disclosures were shared by participants and discussed during the interviews. Our analysis identified four themes: A spectrum of AI uses; Multi-factorial disclosure decisions; Discrepancies between actual and disclosed use; and Emotions around disclosure. These are described below and illustrated with representative excerpts from the dataset.

**Discussion:** AI-disclosure practices take place at the intersection of shared ethical principles and situated pragmatic concerns. Disclosure is both an intellectual and an emotional practice, changing over time as AI use evolves and journal expectations develop. More generative uses may provoke divisive responses and are related to uncertainty about how to disclose, concern about disclosure penalties, and periodic use-disclosure discrepancies. Understanding how researchers are navigating these complexities may help our HPE community articulate clearer guidance for transparent and meaningful disclosure of AI-use in scholarly work. Reflective methodologies can help bring these issues to the surface of our field’s ongoing conversations about AI use.

## INTRODUCTION

AI use is growing by scholars, and disclosure of that use is required by scientific ethics and journal policy. However, AI-use disclosure is emerging as a fraught practice. There is growing unease about the meaning, and meaningfulness, of AI-use disclosures in published manuscripts and little is known about when and how authors decide to represent their AI use in their disclosures. Without such understanding, we are ill-equipped to redirect disclosure practices for increased sufficiency and sincerity, which undermines our efforts towards transparency in the era of AI-supported scholarship.

AI disclosure is a widely accepted principle but a somewhat murky practice. Publisher and journal policies are clear that AI use should be disclosed^1,2^, but they tend to frame AI use more negatively than positively^3^ and they provide inconsistent and imprecise guidance about when and how AI uses should be disclosed. For instance, *Medical Education’s* AI policy for authors states that “We do not mandate disclosure of AI usage in every instance…, [however] authors should disclose AI usage in their manuscript when it is fundamental to their research”^4^, which leaves the judgment of “fundamental” use up to the authors. This can be problematic because, while scholars support transparency in principle, survey results show that they disagree about the necessity of disclosure in particular circumstances.^5^ Varied perceptions of disclosure appear to reflect several factors, including uncertain disclosure thresholds^6^, quickly evolving guidance from journals^7,8,^ and disciplinary differences. Some scholars challenge the principle of transparency itself on the basis that, because other forms of technical support (e.g., spell check) and creative inspiration (e.g., art) do not require disclosure.

Objectively, disclosure lags behind use. Health professions education (HPE) journal editors report seeing very few disclosures in 2024-2025,^6^ and bibliometric analysis suggests that HPE follows the pattern of other scholarly disciplines^9^ with only 2.5% of published empirical manuscripts in the first half of 2025 containing an AI-use disclosure statement.^10^ Overall, scholars tracking the relationship between AI use and disclosure raise concerns related to a “hidden ecosystem of AI use”^11^. When AI use *is* disclosed in published manuscripts, concerns have been raised about the quality and meaningfulness of those disclosures, which may signal compliance rather than transparency.^12^ In HPE, for example, a practice of ‘safe disclosure’ has been noted, in which authors acknowledge AI use in ways that minimize perceived risk or scrutiny.^10^ Such practices may reflect the transparency dilemma in which authors are encouraged to disclose AI use, yet disclosure itself can invite criticism, stigma, or questions about the legitimacy of the work, potentially encouraging authors to disclose minimally or selectively.^13^ A recent HPE commentary highlights this tension, describing an instance in which authors who transparently disclosed AI use were criticized by peer reviewers for using AI.^14^ When such implicit disclosure penalties occur, they can create reluctance to disclose.^15^

Against this backdrop, disclosure frameworks and guidelines are an attempt to improve disclosure quality.^16,17,18^ However, such efforts cannot succeed without better insight into the relationship between AI use and AI disclosure in specific situations in HPE. Responding to calls for empirical research into “when and why authors choose to disclose (or not), how they make decisions about what details to include, and whether their disclosures accurately describe how AI factored into their workflow”,^16^ this study asked two research questions: How do HPE scholars describe their decisions about whether, what, and how to disclose their use of AI in scholarly work? And, what factors shape those disclosure decisions? Close study of these decisions is critical if we hope to understand how the ethics of transparency and the pragmatics of disclosure intersect in daily scholarly practice.

## METHODS

Using descriptive qualitative research methodology^19^, we conducted semi-structured interviews with participants who publish in HPE journals as first authors. The study received ethical approval from Western University (ID#125269).

Sampling was purposive and iterative. We began by inviting authors identified in a bibliometric study of HPE manuscripts with AI-use disclosure statements published in the first six months of 2025^10^; 7 authors (of 50 invited) participated. Realizing that disclosure practices are fast evolving, we then invited authors with papers published in the first 6 months of 2026 from two HPE journals, *Perspectives on Medical Education* and *Academic Medicine*, to which co-authors had editorial access to metadata indicating presence of an AI disclosure; 11 (of 24 invited) authors participated. In both recruitment phases, we reviewed the published disclosures prior to inviting participants, in order to ensure a variety of disclosure types in terms of level of detail and types of AI use described. We also sought to include a range of authors across career stage, geographic location, and research approach. Recruitment continued until iterative analysis suggested data adequacy had been achieved, a judgment based on analytical depth rather than sample size and requiring themes to be well-described and their relationships clearly characterized^20^. In total, we interviewed 18 participants. They included 13 women and 5 men and represented 7 countries (Australia, Canada, Netherlands, New Zealand, Singapore, Thailand, USA), early (almost completed PhD candidates), mid (associate professor) and late (full professor) career stages, and qualitative and quantitative research approaches

Interviews were conducted by Zoom and lasted 40-60 minutes. The interview questions covered five domains: Use of AI, Decision to disclose, Writing the disclosure, Team discussions, Policy guidance, Emotions about disclosure (See Supplement 1 for interview guide). For participants who shared multiple published disclosures, we cycled through these domains again as relevant. The authors each conducted a subset of the interviews. HPE is a relatively small and integrated field, so we assigned interviewers thoughtfully to strengthen rapport and minimize conflicts; for instance, we did not interview our own institutional colleagues but did, in some cases, interview our national and international collaborators. We also considered how our professional roles and prior experiences might shape data collection and interpretation. LL, LM, and ED are senior

HPE researchers and journal editors whose scholarship includes AI use and disclosure in academic publishing; their familiarity with the topic informed their interview conversations and was explicitly considered during team discussions of the developing analysis. MA is a medical student who is familiar with his university’s AI-use policies and has contributed to a previous research project exploring AI disclosure in HPE.

Recordings were initially auto-transcribed by Zoom (institutional account) and manually checked for accuracy and anonymized by MA. The anonymized transcripts were analyzed using reflexive thematic analysis (Braun and Clarke 2008, 2021) to develop themes inductively from the interview data. Following the first few interviews, MA circulated the transcripts to the research team and facilitated a discussion of early codes and further lines of questioning to explore. As more interviews were conducted, each team member shared their reflective interview notes to support email discussion of new patterns and sampling strategies to explore them. As new transcripts were added to the analysis, LL and MA held regular discussions to review the codes and identify early themes. MA used generative AI to assist in tracing overlaps among codes and tighten code definition language to showcase distinctions and relationships. As the final few transcripts were added, LL led the final phases of analysis and interpretation because MA’s summer research placement was ending. These final phases included research team meetings via Zoom to discuss potential overarching themes, as well as substantive written feedback on early drafts of thematic results to refine the key contributions of the study and discern whether these warranted one manuscript or two.

## RESULTS

Participants reported a wide range of experience writing AI-use disclosures (from 1-16 disclosures/participant); in total 31 published disclosures were shared by participants and discussed during the interviews. Our analysis identified four themes: A spectrum of AI uses; Multi-factorial disclosure decisions; Evolving and discrepant disclosure practices; and Emotions around disclosure. These are described below and illustrated with representative excerpts from the dataset.

### A Spectrum of AI uses

Participants’ decisions about AI disclosure were closely tied to how they understood and characterized their own AI use. They described using AI for a variety of purposes, ranging from more assistive uses like editing to more generative uses like “sparring partner” (P17). All participants described the assistive end of this spectrum, expressing familiarity and comfort with technical uses such as “improve writing I’ve already produced” (P12), “revise and polish” (P7), or “reduce this by 50 words” (P4). Similarly, all participants explicitly invoked the principle of human oversight of AI-generated content, affirming that “responsibility still has to lie with the authors” (P2).

Many participants who discussed their AI use over time and referenced multiple disclosures traced a shift in their use from assistive to more generative tasks. One characterized their use as having “evolved from a very basic use of the tool, like drafting an abstract or fixing paragraph, fixing sentences, to this idea of brainstorming. And a stress test.” (P14) Another explained, “I’ve moved from more grammar, stylistic, [uses] …to having it also challenge me and providing feedback” P16).

Participants who had made this shift characterized it positively. Expressing excitement, one called it “a fab tool”, “fascinating”, explaining that “I would never have been able to understand that topic if I hadn’t had a chat with Chatty [ChatGPT] about Foucault and how Foucault might approach different things within my discipline” (P6). The shift to more generative uses was also narrated as a maturing of their own AI skills: “waking up” to the potential of AI was how one participant referred to their shift from using AI as a coding tool to using it for “thinking about the framing and the story” (P13).

However, generative uses of AI provoked divisive responses from other participants. Some viewed generative uses as a source of concern and shared their own sense of boundaries of acceptable AI use, particularly around using AI for qualitative analysis and idea generation. Regarding the former, a participant explained that “qualitative research depends on interpretation and reflexivity… That’s why I’m comfortable using AI for structured tasks but not for drawing conclusions…”. (P12) Regarding idea generation, some participants shared their sense that original ideas should remain the provenance of the human researcher. Scholarly idea generation was described as “high stakes”: “I would consider the writing of a manuscript or a written scholarly piece, that would, for me, be more… more high stakes, and then I would like to be more in control of the initial coming into existence of the idea” (P17). Idea generation was viewed as related to “academic integrity [which] requires me to come up with the ideas and the thoughts and the reasoning myself. I’m okay to use an AI for my abstract, or to cut a hundred words, or those sorts of things, but the argument, the manuscript, the research, all of that better be me.” (P8) These differences in how participants described using AI, whether as assistive or generative tools, shaped how they thought about disclosure, including whether it was needed and how much detail was sufficient.

### Multi-factorial disclosure decisions

Participants described disclosure as a series of decisions about whether AI use warranted disclosure, what aspects of that use to include, and how specifically to describe it in their disclosure statement. Their decisions were shaped by how they conceptualized the nature of their AI use, available journal guidance, concerns about transparency, and anticipated reactions from peer reviewers and editors.

Disclosure was widely viewed as necessary, with advice to “just disclose it always” (P5) as a matter of “academic honesty” (P9), in part due to the sense that “being vague can be a form of being not fully truthful” (P15). Nevertheless, non-specific disclosures were commonly described. One participant shared, “usually it’s a pretty broad kind of motherhood statement. It’s not … go into the depths of… how you use it.” (P5) Another characterized their disclosures as “kind of a standard basic statement and not revolutionary” yet acknowledged “but I think that would be a gross underrepresentation of the actual process of what happens.” (P10). Other participants argued for specifics, reasoning that “journals care about *why* you used it, not just that you used it”:

> “using AI to organize information or improve writing clarity is very different from asking it to interpret findings or develop the intellectual argument of a manuscript. Those different uses have different ethical implications, and our disclosure practices should reflect those differences.” (P8)

How, exactly, disclosure practices should reflect these different uses was less clear in our data. Participants routinely referred to journal policies to guide disclosure practices, yet they found that journals were “still feeling their way” (P1): “more support and guidance” (P4) from journals was a widely expressed need.

Participants expressed uncertainty about how to word disclosures. Even seemingly straightforward journal guidance could be difficult for the participants to interpret. For example, one participant questioned: “you must declare if you used AI for anything more than grammar and light proofing. It’s like, well, what’s light proofing? My light proofing is probably different to an undergrad student who’s never written before. So, what is light proofing?” (P2) This uncertainty was even more pronounced in generative AI use cases. For instance, in reference to brainstorming with AI early in their research process, one participant wondered, “Is that something that should be declared?” (P18) Another asked: “if I went on and wrote a manuscript about that [AI chat session about Foucauldian takes on their ideas], should I disclose that I had my ideas about the subject influenced by ChatGPT?” (P6) Some participants acknowledged that the projects in which they’d used AI more generatively were not yet at the submission stage, and weren’t yet sure how they intended to handle the disclosure:

> “I’ve used AI to brainstorm ideas by asking questions like, ‘Here’s an idea. Help me think through an appropriate study design.’ I haven’t yet published a study that relied on that kind of interaction, but I do wonder how I would disclose it. I think that’s an interesting question. I’m not sure what the appropriate practice should be” (P12).

Grappling with uncertainty about disclosure during the interview, participants returned to the principle of transparency. They reported that “I disclose everything that I used it for” and characterized their disclosures as “going absolutely above and beyond what I thought any reasonable person might want to know” (P3). Some also felt it necessary to write negative disclosures stating that AI was *not* used for more generative tasks, such as “these tools were used exclusively to improve clarity of sentence structure and address grammatical nuances. The authors did not use generative AI for ideation, conceptual development, or drafting of content at all stages” (P11). When asked why, this participant reflected “if you said that you’ve used AI for ideation and then all of that, then there will be more questions from either the reviewer or the, um, or the editor” (P11).

### Discrepant disclosure practices

As participants shared and reflected on multiple disclosures from the past months, some interviews surfaced discrepancies between the AI use that participants described to the interviewer and what appeared in their published disclosures. For instance, consider the following exchange mid-interview with a senior HPE scholar:

Interviewer: Okay, so let me probe this just a little bit more and be a bit provocative. Why doesn’t your disclosure say [what you just described to me], that ‘this whole concept comes from my first conversation with AI about this’ … [your published disclosure] is pretty generic. I presume that was a decision.

P15: Probably. Or a non-decision, I don’t know. Yeah. Again, I probably and I still feel whenever I use AI and I and I come up with something that I think is good with its help as a writing partner I still have this little twinge in my stomach of this wouldn’t have happened without the help of this tool, and so is it really fully mine? And so maybe, maybe I was not comfortable sharing … those specifics, yeah. I don’t know, don’t I don’t remember, consciously saying, I’m not going to say that I came up with the model that way, but yeah, it’s a good point. Like, I probably should have.

Another participant worried she couldn’t attest to the accuracy of her published disclosure: after reviewing it with the interviewer, she admitted “I’m suddenly struck that maybe I don’t know exactly how much it was used” (P3). These discrepancies don’t necessarily represent a deliberate decision to withhold information. Many participants only recognized the discrepancy when invited to reflect on the disclosure during the interview. One recognized that “the description that I gave you about the [AI-produced] figure, isn’t…doesn’t come through in the disclosure that we wrote. And so, it might have been useful for the audience to understand a little bit more about how AI impacted the development of the figure” (P4). Reflecting on the discrepancy, they explained that the decision to keep the published disclosure minimal “came from two places. It came from the vulnerability, feeling of vulnerability, and how other people are going to interpret what this is. And then also, just going back to the word count issue, honestly” (P4).

Participants particularly grappled with transparency when their AI-use was perceived to be more substantive: “that’s a struggle of, like, how do we stay truthful and honest, and we have to have a minimum bar, but, you know, again, it’s degrees of, like, how much do we explain how heavily we relied, or do we use vagaries to kind of feel comfortable?” (P13)

### Emotions around disclosure

Disclosure was experienced as not only an intellectual practice but also an emotional one. One participant shared that “the ultimate decision [to disclose] wasn’t actually that hard, but I had to do my internal, like, emotional processing, I think, before I did it” (P10). Participants shared that they felt “a little nervous” (P2), “scared” (P7), “anxious” (P15) “some fear, a little vulnerability” (P4). Those emotions resurfaced for some participants while reviewing their disclosure during the interview: “I can feel my own… my previous emotions, like, coming up around. Yeah, I get … this feeling that it’s cheating somehow, … that it needs to be, like, surveilled.” (P10)

Emotions were portrayed as becoming more positive over time. Authors comparing their 2025 disclosure experience with current practice said, “I’m much more familiar and comfortable with them now probably than I was then” (P2) and “I’m now less hesitant, or I’m not scared anymore when I’m the first author of the… of a manuscript, to immediately put [the disclosure] in” (P17). The field’s growing uptake of AI was a factor in this emotional evolution: “I feel less nervous about it now because I think it is so pervasive” (P2). However, not everyone was feeling more at ease: for instance, this participant shared his persistent worry about “people looking at you funny. I still feel that way. ‘He didn’t write that, he had help’” (P15). There was a shared sense that disclosure “opens up a can of worms” (P9); one author characterized the feeling as being “the guinea pig that gets experimented on with this” as “there’s still no clarity” about how universities and journals will respond (P1).

Personal experience and career stage influenced emotions. One senior researcher asserted “I don’t feel guilty and I don’t feel embarrassed” (P8) about disclosing AI use; others noted that “the first few times I might’ve been worried of the backlash it could have brought, but I think that has shifted at least for me. And it also, I guess it makes the, yeah, the place I am in my career makes a difference with how I approach this” (P16). By contrast, some early career participants had strategically avoided any AI use and disclosure until they felt more secure in their identity: “I did not want to run a risk at this late stage in my thesis, being almost at the finish line to start tinkering with anything that could potentially bring me into question, so I’ve chosen to actively avoid it” (P1).

Their supervisors were aware, with comments on the “angst” (P3) their graduate students were feeling about disclosure.

## DISCUSSION

Our findings characterize AI-use disclosure in HPE as a contested, evolving, multi-factorial and emotional practice. These are experiences of HPE researchers who are already using and disclosing generative AI, giving us a sense of the ‘front line’ of AI disclosure experience that is helpful as our field develops its shared understanding of the role of AI in HPE scholarship. Below we discuss our four main themes and place them into conversation with existing literature on AI disclosure.

Four years after its appearance, generative AI use remains a contested subject, even among HPE scholars experienced with AI-use disclosure. AI is being used for both assistive and generative tasks, with an apparent shift towards the latter that resonates with results of a recent survey on AI uses by the public in 2026.^21^ Our participants framed assistive tasks like editing and translation matter-of-factly and did not portray them as challenging to disclose. However, as has also been reported in other fields^22,23^, more generative uses provoked divisive responses. Some of our participants viewed the use of AI for brainstorming and conceptualization as a threat to scholarly identity and integrity, while others described these as their most exciting AI interactions. This divide was at least in part related to research approach: qualitative participants expressed more reservation about using AI for generative uses during analysis and interpretation than quantitative participants did. Their reservations are situated in a larger public debate in which qualitative researchers from 32 countries have rejected the use of AI for qualitative thematic analysis due to fundamental methodological incongruence.^24^ It appears that, as a highly interdisciplinary field, HPE is experiencing within its borders the “substantial disciplinary variation” that comparative research across fields has identified in scholars’ perceptions of appropriate AI use^25^. We have worked hard over the past 20 years to bridge methodological approaches and respect paradigmatic differences in HPE research^26,27^; our results suggest that AI may inflame anew these interdisciplinary flashpoints we had believed largely resolved^28^. Recognizing this, we must give them the attention they deserve as we create guidelines and policies to support AI-use disclosure.

While participants all agreed in theory that disclosure was necessary for transparency and scientific integrity, they described their specific disclosure processes as complex and multi-factorial. The lack of consistency and clarity in journal guidelines was widely perceived as a barrier, and ambiguous language such as “light proofing” or “fundamental” contribution did not help participants judge the necessity threshold where disclosure would be expected.^6^ More generative uses were particularly problematic: many were uncertain about how to describe these uses and worried about how peers and journal editors would respond to such disclosures. We wonder if this is giving rise to negative disclosures specifying that authors have *not* used AI for such uses as conceptualization and ideation.^10^ Given that more generative uses seem to be increasing in our field and others^20,21^, we worry that this may intensify an apparent gap between AI-use and AI disclosure in HPE.^10^

Clearer policy and guidelines are one way to close this gap, and a recent AMEE guide signals progress in this direction.^16^ However, we agree with Yoon et al that “disclosure practices are shaped not only by formal publication requirements but also by localized understandings of what constitutes responsible and legitimate research conduct” (p12).^14^ Policy is necessary but insufficient; we would contend that candid and reflective community discussion is also required. To make a difference, though, such discussion must move beyond abstract principles: what we need to understand is the nuances of how the ethics of transparency and the pragmatics of disclosure intersect in daily scholarly practice. Our methodology had this as its aim; it is the first study to our knowledge to ask scholars to reflect on their own, specific, published AI-use disclosures. This approach produced rich and emotive conversations as participants explained their AI-use in a project, compared that use to their published disclosure, and reflected on any discrepancies either they or the interviewer noted. While discrepancies were not frequent in our data, those identified shared some key characteristics: they were generically accurate but lacked detail regarding exactly how AI was used, particularly when uses were more fundamental, such as AI suggesting the central concept underpinning the work. Further, participants’ reflections suggest that such discrepancies may be rhetorically (if perhaps unconsciously) motivated by fears of social stigma associated with more fundamental uses. These preliminary discrepancy results resonate with bibliometric evidence that HPE disclosures are “safe”: brief and generic descriptions of use that are curated to protect against perceived risk.^10^ We believe that disclosure discrepancies require further research, because, until we understand them more fully, the ubiquitous objective of “increased transparency”^29^ remains out of reach.

Our results also highlight the emotional aspects of AI-use disclosure, introducing a subtle shift in the current scholarly conversation. Policies and guidelines have predominantly treated disclosure as a rational practice motivated by academic standards for integrity and transparency. Even when emotions are noted, they are dismissed rather than engaged: e.g., our AMEE Guide recommended “that authors approach disclosure as reporting rather than confessing. AI-use is not something to be ashamed of” (p548).^17^ The recent literature on disclosure penalties are beginning to shift this instrumental (“just do it”) disclosure conversation towards a more critical one.^12, 13^ Our work continues this shift by describing how HPE scholars *feel* about disclosure: often afraid, vulnerable, ashamed, and worried. These results resonate with recent descriptions of the “moral shame” of AI-disclosure^30,31^ in other fields. Further, a project to measure AI-users’ “shame and guilt” posits a connection between these emotions and imposter syndrome^32^, which may be a useful lens for understanding our early career participants’ view of disclosure as a scary proposition until they acquired sufficient credibility in the field that readers would know they could have done the work without AI’s assistance. Finally, while our participants believed that emotions are shifting as our field becomes familiar and disclosure is normalized, the prominence of negative emotions in our dataset suggests that we should not assume that they will dissipate easily, or evenly, across the HPE field. We encourage further study of disclosure as an emotional practice, with particular attention to how disempowered, marginalized and multi-lingual scholars are experiencing and responding to the affective threat of disclosure penalty.

### Limitations

While purposive, our sample was also based on convenience, and it may be that those who did not respond to our invitation have different views and experiences than our participants. For instance, their non-response could suggest less interest in AI-use disclosure, or greater vulnerability and fear than authors we spoke with. Readers will need to judge transferability of our findings by considering both what our study sample includes and what it overlooks.

Our insight into use-disclosure discrepancies is limited by two features of our design. First, our interviews with participants were many months after their AI-disclosure was written, and many more months after their AI-use behaviors, introducing the challenge of hindsight bias. Second, a close, comparative linguistic analysis of both the published disclosures and researchers’ descriptions of their AI use was beyond the scope of the current study, which means that our insights are preliminary and further work is needed to characterize use-disclosure discrepancies in more granular detail.

## CONCLUSIONS

AI-disclosure practices take place at the intersection of shared ethical principles and situated pragmatic concerns. This gives rise to strong emotions, multi-factorial disclosure decisions, and periodic use-disclosure discrepancies. HPE needs to acknowledge that there is a spectrum of AI uses, because the pragmatics of disclosure appear to vary across this spectrum. Reflective methodologies can help bring these issues to the surface of our field’s ongoing conversations about AI use.

## Supporting information

Supplement 1_Interview Guide

## Data Availability

To protect participant anonymity, transcripts are not available upon request.

## Acknowledgements

*This project was funded by a 2024 Digital Research & Innovation Grant from Continuing Professional Development, Schulich School of Medicine & Dentistry, Western University*.

## The authors have no conflicts of interest to declare

*ChatGPT 5.0 was used after initial coding of the initial 7 transcripts to help identify areas of overlap among the author-generated preliminary codes, and to refine the wording of preliminary code labels. All suggestions were discussed during subsequent analysis meetings, to verify their relevance and accuracy, and to situate them within the team’s developing understanding as new transcripts were added to the analysis. ChatGPT was not used during later stages of analysis as themes were developed and finalized, and interpretations were negotiated with reference to current literature on disclosure. The authors take full responsibility for all material presented in the manuscript*.

## References

1 COPE Council. COPE position - Authorship and AI - English. (2024) 10.24318/cCVRZBms Committee on Publication Ethics. Accessed September 21, 2026.

2 Mary Beth DeVilbiss, Laura Weiss Roberts, Artificial Intelligence Tools in Scholarly Publishing: Guidance for Academic Medicine Authors, Academic Medicine, Volume 98, Issue 8, August 2023, Pages 865–866, 10.1097/ACM.0000000000005261

3 Saravanan S, Chan SC, Rashid MA. Artificial intelligence in health professions education publishing: A content analysis of journal and publisher guidance. Medical Teacher. 2026 Aug 20:1–6.

4 Medical Education, Guidelines for Authors: AI Policy. Accessed September 21, 2026. https://asmepublications.onlinelibrary.wiley.com/hub/journal/13652923/homepage/author-guidelines

5 Kwon D. Is it OK for AI to write science papers? Nature survey shows researchers are split. Nature. May 2025.

6 Lingard L, Driessen E, Oswald K. The blurred threshold of AI-use disclosure: International journal editors’ expectations of sufficiency and necessity; 2025. DOI: 10.1101/2025.07.17.25331725

7 Flanagin A, Pirracchio R, Khera R, Berkwits M, Hswen Y, Bibbins-Domingo K. Reporting Use of AI in Research and Scholarly Publication. Network Guidance. JAMA. 2024;331(13):1096–1098. DOI: 10.1001/jama.2024.3471

8 Flanagin A, Perlis RH, Bibbins-Domingo K. Updated Guidance for Author Use of AI in Medical Publication. JAMA. 2026;336(11):926–928. doi:10.1001/jama.2026.16613

9 Jonah Barrett D, Heng R, Perchik JD. Documenting Disclosure: Limited Reporting of Generative AI Usage in Radiology Research Manuscripts. Acad Radiol. 2025 Oct;32(10):5686–5692.

10 Ans M, Maggio LA, Algodi H, et al. The presence and nature of AI-use disclosure statements in medical education journals: A bibliometric study. Perspect Med Educ. 2026;15(1):212–225. doi: 10.5334/pme.2431

11 Fiorillo L. Confronting the demonization of AI writing: reevaluating its role in upholding scientific integrity. Oral Oncol Rep. 2024;12:100685. doi: 10.1016/j.oor.2024.100685

12 Hartberg Y. AI Disclosures are A Joke. Chronicle of Higher Education. August 2025. Accessed online Sept 12, 2026.https://www.chronicle.com/article/ai-writing-disclosures-are-a-joke-heres-how-to-improve-them.

13 Schilke, O., & Reimann, M. (2025). The transparency dilemma: How AI disclosure erodes trust. Organizational Behavior and Human Decision Processes, 188, 104405.

14 Maggio, L. A. (2026). Mixed signals: When AI disclosure becomes a liability. Medical Teacher, 1–3.

15 Yoon, A., & Oristaglio, S. (2026). When AI use becomes the norm: Researcher perspectives on AI disclosure policy and practice. Accountability in Research, 2716884.

16 Weaver KD. The AI Disclosure (AID) Framework. C&RL News. 2024;85(10):407–411.

17 Cleland J, Driessen E, Masters K, Lingard L, Maggio LA. When and how to disclose AI use in academic publishing:AMEE Guide No.192. Medical Teacher. 2025;1–12. DOI: 10.1080/0142159X.2025.2607513

18 Cheng A, Calhoun A, Reedy G. Artificial intelligence-assisted academic writing: recommendations for ethical use Advances in Simulation. 2025;10:22.

19 Chafe R. (2017). The value of qualitative description in health services and policy research. Healthcare Policy, 12(3), 12. https://www.ncbi.nlm.nih.gov/pmc/articles/PMC5344360/

20 Braun, V., & Clarke, V. (2022b). Thematic analysis: A practical guide. Sage.

21 Sanders MZ. (2026) Harvard Business Review https://hbr.org/2026/06/how-people-are-really-using-ai-in-2026 Accessed Sept 8, 2026.

22 Anderson JP, Degn L, Fisherg R, Graversen EK, Horbach SPJM, et al. (2025) Generative Artificial Intelligence (GenAI) in the research process – A survey of researchers’ practices and perceptions. Technology in Society, Volume 81.

23 Ma R, Dedema M, Cox A. (2025) A dancing bear, a colleague, or a sharpened toolbox? The cautious adoption of generative artificial intelligence technologies in digital humanities research. DOI: 10.48550/arXiv.2404.12458.

24 Jowsey, T., Braun, V., Clarke, V., Lupton, D., & Fine, M. (2025). We Reject the Use of Generative Artificial Intelligence for Reflexive Qualitative Research. Qualitative Inquiry, 0(0). 10.1177/10778004251401851

25 Ayoung Yoon & Siena Oristaglio (13 Aug 2026): When AI use becomes the norm: Researcher perspectives on AI disclosure policy and practice. Accountability in Research, DOI: 10.1080/08989621.2026.2716884

26 Georges Bordage, Moving the Field Forward: Going Beyond Quantitative–Qualitative, Academic Medicine, Volume 82, Issue Supplement_1, October 2007, Pages S126–S128, 10.1097/ACM.0b013e31813e661d

27 Eva, K.W. (2009), Broadening the debate about quality in medical education research. Medical Education, 43: 294–296. 10.1111/j.1365-2923.2009.03342.x

28 Cleland J, Masters K, Driessen E, Kuper A, Change YC, Lingard L. “The Death of the Academic Author? Implications of Artificial Intelligence for academic writing and publishing”. AMEE 2026 Symposium, August 26, 2026, Vienna Austria.

29 Government of Canada. Enhancing trust in artificial intelligence through increased transparency. https://ised-isde.canada.ca/site/ised/en/have-your-say-advancing-ai-transparency-canada/enhancing-trust-artificial-intelligence-through-increased-transparency Accessed September 21, 2026.

30 Bao, A., & Zeng, Y. (2026). AI disclosure, moral shame, and the punishment of honesty. Accountability in Research, 33(3). 10.1080/08989621.2025.2542197

31 Acut, D. P., Gamusa, E. V., Pernaa, J., Yuenyong, C., Pantaleon, A. T., Espina, R. C., Sim, M. J. C., & Garcia, M. B. (2025). AI shaming among teacher education students: A reflection on acceptance and identity in the age of generative tools. In Pitfalls of AI integration in education: Skill obsolescence, misuse, and bias (pp. 95–118). IGI Global Scientific Publishing.

32 Cipriani, E., Menicucci, D., Greco, A., & Grassini, S. (2026). Emotional Responses to AI Use: Development of the SAG-AI Scale for Shame and Guilt. International Journal of Human–Computer Interaction, 1–20. 10.1080/10447318.2026.2648803

