## Supplement 1_Interview Guide for "A “vulnerable” space: Health professions education scholars’ reflections on AI-use disclosure"

**Interview guide for authors of published manuscripts with AI-use disclosure statements**

Background and Context

- Could you tell me a bit about your professional background and research area?
- How familiar were you with using generative AI tools before working on this manuscript?

Use of genAI in the Manuscript

- Please describe how you used genAI during the development of the paper we identified.
- What motivated you to use genAI in this context?
- Were there any aspects of writing, analysis, or communication where genAI was particularly helpful or challenging?

Decision to Disclose

- How did you decide whether to disclose your use of genAI in the paper?
- What factors most influenced that decision (e.g., journal policy, co-authors, ethics, perceptions of reviewers)?
- Did you look at examples of other disclosures or consult any guidance? Please describe.

Process of Drafting the Disclosure

- How did your author team determine what details to include in the disclosure?
- Were there aspects of genAI use that you chose not to include? Why?
- How easy or difficult was it to decide on the wording or placement of the disclosure?

Team Dynamics and Discussion

- Did the decision to disclose come up in discussions with your co-authors?
- Were there differing opinions among the team? If so, how were those resolved?
- Did any institutional or supervisory perspectives shape those discussions?

Perceptions of Policy and Practice

- How clear were the journal’s expectations about genAI use and disclosure?
- Did you have any contact with editors about your disclosure before or during submission?

Emotional and Ethical Dimensions

- How did you feel about disclosing your AI use? (e.g., confident, anxious)
- What ethical or professional considerations factored into your decision?

Reactions and Outcomes

- How did reviewers or editors respond to your disclosure, if at all?
- How, if at all, did your disclosure seem to shape the review or editorial decision process?
- Can you describe any other experiences you’ve had disclosing genAI use in manuscripts? How many disclosures would you estimate you’ve included in submitted manuscripts? How have those earlier disclosures influenced the way you approach disclosure now?

Closing

- Is there anything else you would like to share about your experience disclosing genAI?
- Do you have any questions for me about the study?
